# A Simple RT-PCR Melting temperature Assay to Rapidly Screen for Widely Circulating SARS-CoV-2 Variants

**DOI:** 10.1101/2021.03.05.21252709

**Authors:** Padmapriya Banada, Raquel Green, Sukalyani Banik, Abby Chopoorian, Deanna Streck, Robert Jones, Soumitesh Chakravorty, David Alland

## Abstract

**Background:** The increased transmission of SARS-CoV-2 variants of concern (VOC) which originated in the United Kingdom (B.1.1.7), South Africa (B1.351), Brazil (P.1) and in United States (B.1.427/429) requires a vigorous public health response, including real time strain surveillance on a global scale. Although genome sequencing is the gold standard for identifying these VOCs, it is time consuming and expensive. Here, we describe a simple, rapid and high-throughput reverse-transcriptase PCR (RT-PCR) melting temperature (Tm) screening assay that identifies these three major VOCs.

**Methods:** RT-PCR primers and four sloppy molecular beacon (SMB) probes were designed to amplify and detect the SARS-CoV-2 N501Y (A23063T) and E484K (G23012A) mutations and their corresponding wild type sequences. After RT-PCR, the VOCs were identified by a characteristic Tm of each SMB. Assay optimization and testing was performed with RNA from SARS-CoV-2 USA WA1/2020 (WT), a B.1.17 and a B.1.351 variant strains. The assay was then validated using clinical samples.

**Results:** The limit of detection (LOD) for both the WT and variants was 4 and 10 genomic copies/reaction for the 501 and 484 codon assays, respectively. The assay was 100% sensitive and 100% specific for identifying the N501Y and E484K mutations in cultured virus and in clinical samples as confirmed by Sanger sequencing.

**Conclusion:** We have developed an RT-PCR melt screening test for the three major VOCs which can be used to rapidly screen large numbers of patient samples providing an early warning for the emergence of these variants and a simple way to track their spread.

## Introduction

In December 2020, public health officials in the United Kingdom observed a surge of COVID-19 cases in Kent, England that appeared to be largely due to a specific variant of SARS-CoV-2 (1, 2). The new variant was named Variant of Concern (VOC 202012/1) or B.1.1.7 based on its phylogenetic lineage. Early reports suggested that B.1.1.7 was more transmissible and possibly more virulent than previous SARS-CoV-2 strains (3, 4), although the evidence did not suggest that B.1.1.7 caused a decrease in vaccine efficacy (5-8). The B.1.1.7 variant has a number of mutations in the spike protein including single nucleotide polymorphisms (SNPs) resulting in N501Y, A570D, D614G and P681H mutations, and deletions at amino acids 69-70 and 144Y (6, 9). The N501Y (A23063T) mutation has been identified as an important contributor to the worrisome phenotype (2, 4, 10, 11). Other SARS-CoV-2 strain variants have been implicated in large outbreaks in South Africa (known as 20H/501Y.V2 or B.1.351 lineage) and Brazil (20J/501Y.V3 or P.1 lineage) (12). These variants appear to have decreased the efficiency of some COVID-19 vaccines (2, 13, 14). All three, B.1.1.7, B.1.351 and P.1 variants contain the N501Y mutation, while the South African and Brazilian variants additionally contain mutations in E484K and K417N (1, 12). Thus, the N501Y mutation appears to be an excellent marker for all three strains while the E484K mutation can be used to differentiate the other two strains from B.1.1.7. New SARS-CoV-2 variants have also been recently reported in the United States (5, 15). Most of the new variants also have mutations at the 501 and/or 484 position, although additional mutations such L452R have also been reported (15); however, the epidemiological and clinical relavance of these new variants are still not well understood.

All three of the major SARS-CoV-2 N501Y variants are circulating around the world (1). However, the distribution, extent, and spread of these variants are poorly understood in countries such as the United States where only a small fraction (0.04 to 3.5%) of COVID-19 cases are analyzed by viral genomic sequencing as per the CDC’s national genomic surveillance dashboard (16). Viral genome sequencing is currently the only method available to reliably detect rapidly emerging SARS-CoV-2 variants. Genomic sequencing has the advantage of providing a detailed map of new mutations, which supports new variant discovery as well as monitoring the exact type of the variant. However, genomic sequencing is expensive and difficult to perform in real time. In contrast, RT-PCR testing for SARS-CoV-2 has become widespread. This diagnostic approach is easy to perform in a high throughput manner, and rapid turnaround times are possible. However, routine RT-PCR tests do not differentiate among SARS-CoV-2 variants, or do so only by producing negative assay results (17), which still require sequence confirmation (18). Furthermore, the potential for additional mutations to appear near key variant-defining alleles may complicate the development of RT-PCR assays for SARS-CoV-2 variants (19). We have previously demonstrated that sloppy molecular beacons (SMBs) combined with melting temperature (Tm) code analysis may be used to specifically detect mutations in short genomic regions where a variety of mutations can exist (20).

Here, we apply this same Tm-based approach to detect and differentiate variant strains of SARS-CoV-2 with high sensitivity and specificity. This approach is flexible and can be used in high throughput manner, easily allowing the addition of new mutation detecting assays as needed to identify and track new SARS-CoV-2 variants as they emerge. Furthermore, this approach can be performed on a wide range of real-time PCR instrumentation as long as they have the capacity to run melt curve analysis. The wide availability of such instruments can allow quick adoption of this assay around the world, increasing access to real-time monitoring of SARS-CoV-2 variant spread.

## Methods

### Ethical considerations

The use of de-identified clinical samples from confirmed COVID-19 positive and negative patients for PCR testing and sequencing was approved by the Rutgers Institutional Review Board under protocol numbers 20170001218 and 2020001541.

### Viral cultures and RNA

Genomic RNA from SARS-CoV-2 USA WA1/2020 (wild type, WT), viral culture stocks of SARS-CoV-2 hCoV-19/England/204820464/2020 (B.1.1.7 variant, 501-MT) and SARS-CoV-2 Isolate hCoV-19/South Africa/KRISP-K005325/2020 (B.1.351 variant, 484-MT) were obtained from BEI Resources, NIAID (Manassas, VA). RNA was isolated from both the variant strains in a BSL3 laboratory, using RNAdvance viral RNA extraction kit (Beckman Coulter, Indianapolis, IN).

### Genome sequence analysis for assay design

For initial analysis of the mutations in SARS-CoV-2, a total of 330,132 high quality viral genome sequences deposited in GISAID (21) as of Jan 12, 2021, were analyzed. Publicly available datasets were analyzed in this study. This data can be found here: https://www.gisaid.org/. A 250-nucleotide region around the N501Y (A23063T) and E484K (G23012A) positions in the reference strain (GenBank accession number MN908947) was selected and used to identify the corresponding regions in the GISAID dataset using BLAST (22). These matching sequences were condensed into a set of unique sequences and aligned using a multiple sequence alignment program, MAFFT (23). Candidate amplification primers and probes were identified on the basis of sequence conservation and predicted Tm using the algorithm of SantaLucia (24) and final set of primers were designed with the help of the Primer3 program (25). SMB probe design was performed using the web servers DNA mfold (http://www.unafold.org/mfold/applications/dna-folding-form.php) and DINAmelt (http://www.unafold.org/hybrid2.php) to predict the probe folding structures and probe-target hybrid Tm values respectively.

### Primers and Probes

The list of primers and probes used for the 501 variant (SMB-501) and the 484 variant (SMB-484) assays are shown in Table 1. Both assays were run as two separate reactions. For the SMB-501 assay an 89 bp region surrounding the 23063 position was amplified using an asymmetric PCR. For SMB-484 assay, a 76bp region surrounding the 23012 position was amplified using an asymmetric PCR. Sloppy molecular beacons (SMB) were designed targeting both the wild type 501N/484E (23063A/23012A) and the mutant 501Y/484K (23063T/23012G) sequences (SMB-501-MT). Primers were obtained from Millipore Sigma (The Woodlands, TX) and SMBs were synthesized by LGC Biosearch technologies (Petaluma, CA). An internal control (IC) assay developed by CDC (26, 27), targeting the human RNaseP gene was simultaneously performed for each extracted RNA specimen as a separate reaction in a separate well, using the *Taq*Man real-time PCR assay probe tagged with FAM at the 5’ end and Dabcyl quencher at the 3’ end.

**Table 1.**
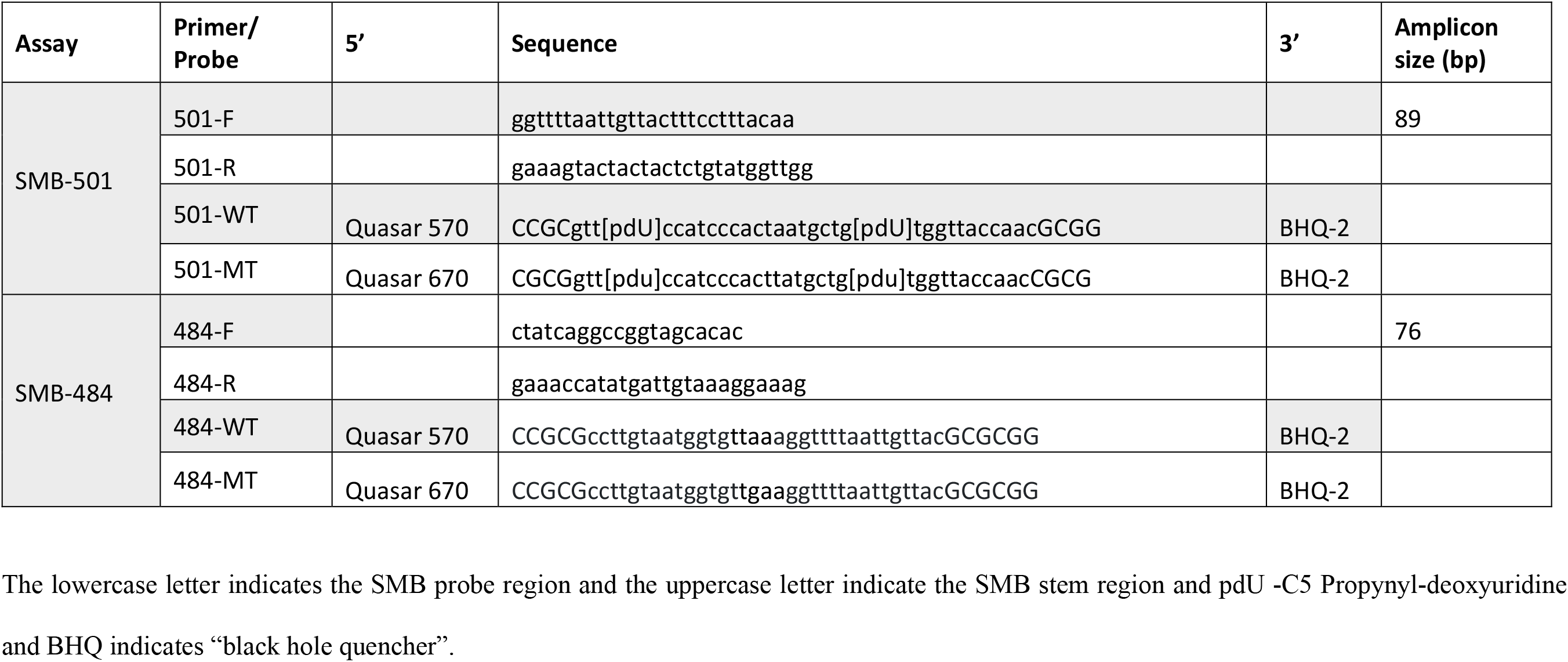
Primers and probes.

### SMB-assay formulation and procedure

TaqPath™ 1-Step RT-qPCR Master Mix, CG (ThermoFisher Scientific, Waltham, MA) was used for the RT-PCR. Each one step reaction mix was supplemented with 0.2µM of the forward primer 501-F/484-F and 2 µM/4 µM of the reverse primer 501-R/484-R, 0.4 µM of each of the SMB probes (SMB-501-WT and SMB-501-MT/ SMB-484-WT and SMB-484-MT) and 1 µl of the template RNA (note that the concentration of the forward primers did not match the concentration of the reverse primers so as to create an asymmetric PCR). The internal control contained primers and a probe specific for human RNaseP as described previously (26, 27). Each reaction was run in replicates of 4 in 384-well plates in a Roche LightCycler 480 (Roche, Indianapolis, IN). The one-step RT-PCR amplification was performed with the following thermocycling conditions: Uracil DNA glucosylase incubation for 2 min at 37°C and reverse transcription (RT) for 15 min at 50°C, followed by asymmetric PCR for 45 cycles (denaturation at 95 °C for 1 s, annealing/extension at 55°C for 30 s). The post-PCR melt was performed with the following conditions: denaturation at 95°C for 30s followed by cooling down to 45°C and gradual heating to 85°C, with continuous monitoring of fluorescence at the rate of 2 acquisitions per °C. The total assay time was 1h 17min. Automated Tm calls were performed by the LC480 Tm detection software at the end of the PCR. The resulting Tm for each probe was identified and matched with the Tm-signature code defined for the wildtype or the mutant variants.

### Analytical sensitivity

The pre-quantitated genomic RNA from the SARS-CoV-2 USA WA1/2020 (WT) obtained from BEI resources with the stock concentration of 1.8 ×10^4^ genomic equivalents (GE)/µl was diluted in Tris-EDTA (TE) buffer. A 1 µl of each concentration (400, 200, 40, 20 and 4 GE/µl for 501 assay and 1000, 500, 100, 50 and 10 for 484 assay) was added to the one-step RT-PCR mix containing the primers and probes and was evaluated in the SMB-501 assay or the SMB-484 assay. The RNA extracted from both variant strains was quantified against a standard curve generated with the N1 gene-specific real time RT-PCR assay (26, 27) and the concentration was determined to be 4×10^5^ GE/µl for B.1.17 and 7×10^6^ GE/µl for B.1.351 variant strain. The mutant RNA was also serially diluted and 1 µl of each concentration mentioned above was evaluated in both the assays.

### Validation with patient samples

Deidentified nasopharyngeal (NP) swabs obtained from patients undergoing routine COVID-19 clinical testing using the Xpert Xpress SARS-CoV-2 or Xpert Xpress SARS-CoV-2/Flu/RSV (4-plex) test (Cepheid, Sunnyvale CA) assay in the CLIA and CAP certified laboratory at the Public Health Research Institute (PHRI), Newark, NJ, were selected for this study. NP swabs were collected in 3ml viral transport media (VTM) from Hardy diagnostics (Santa Maria, CA) or Labscoop (Little Rock, AR) and were banked at either refrigerated conditions (for specimens first tested within the previous 2 weeks) or frozen (for specimens stored longer than 2 weeks). The samples consisted of 46 randomly selected COVID-19 positive samples of unknown genotype and a RT-PCR cycle threshold (Ct) <42 with either Xpert Xpress SARS-CoV-2 or Xpert Xpert CoV-2/Flu/RSV test. Nine of the samples had been banked between October 2020 through December 31, 2020, 16 in the month of January 2021, 15 in February 2021 and 6 in March 2021. Thirty COVID-19 negative specimens were randomly selected from samples banked between Oct 2020 through Feb 2021. RNA was extracted from each specimen using a QiaAmp viral RNA isolation kit, following the manufacturer’s instructions (Qiagen, Valencia, CA) in a BSL2 laboratory. A 5µl volume of RNA was added to the one-step RT-PCR mix containing the primers and probes. A subset of these clinical samples consisting of 26/46 of the COVID-19 positive samples and 19/30 of the COVID-19 negative samples were using the SMB-484 assay, mainly based on the continued availability of the sample for testing. The SMB-501, SMB-484 and the IC assays were tested in separate wells in replicates of two. The samples that failed the IC assay were repeated starting with the RNA extraction. A subset of samples that tested positive either for 501N/484E wildtype or 501Y/484K mutant was confirmed by Sanger sequencing using the primer pair: F-5’ctatcaggccggtagcacac3’ and R-5’ctttcttttgaacttctacatg3’ which amplifies a 143bp segment of the S-gene inclusive of the amino acid positions at 484 and 501. Sequencing chromatograms were analyzed using Ugene (ver 37) comparing against the known WT and MT sequences using MegAlign Pro software (DNAStar, ver16).

### Statistical analysis

Standard statistical analyses (average, standard deviation) and graphing were performed using Microsoft excel (ver 2102) and GraphPad Prism 8.4.3 for Windows.

## Results

### Limit of detection

The one-step RT mix containing asymmetric PCR primers and both wildtype and MT specific probes was added with either WT RNA or MT RNA at different concentrations ranging from 400 to 4 GE/reaction (N=4) for SMB-501 assay and from 1000 to 10 GE/reaction for SMB-484 assay. After Tm analysis was performed the Tm peak heights were highest in the assays with the largest added number of GE and the peak heights progressively decreased as the number of GEs present in the reactions decreased. However, Tm peak heights produced by both SMB 501-WT/ SMB 484-WT and SMB 501-MT/ SMB 484-MT could still be reproducibly detected at the lowest concentration tested (4 or 10 GE/reaction) when tested against both WT and N501Y mutant strains, defining the assay limit of detection as ≤4 GE per reaction for SMB-501 or ≤10 GE/reaction for SMB-484 assay (Fig. 1).

**Fig. 1.**
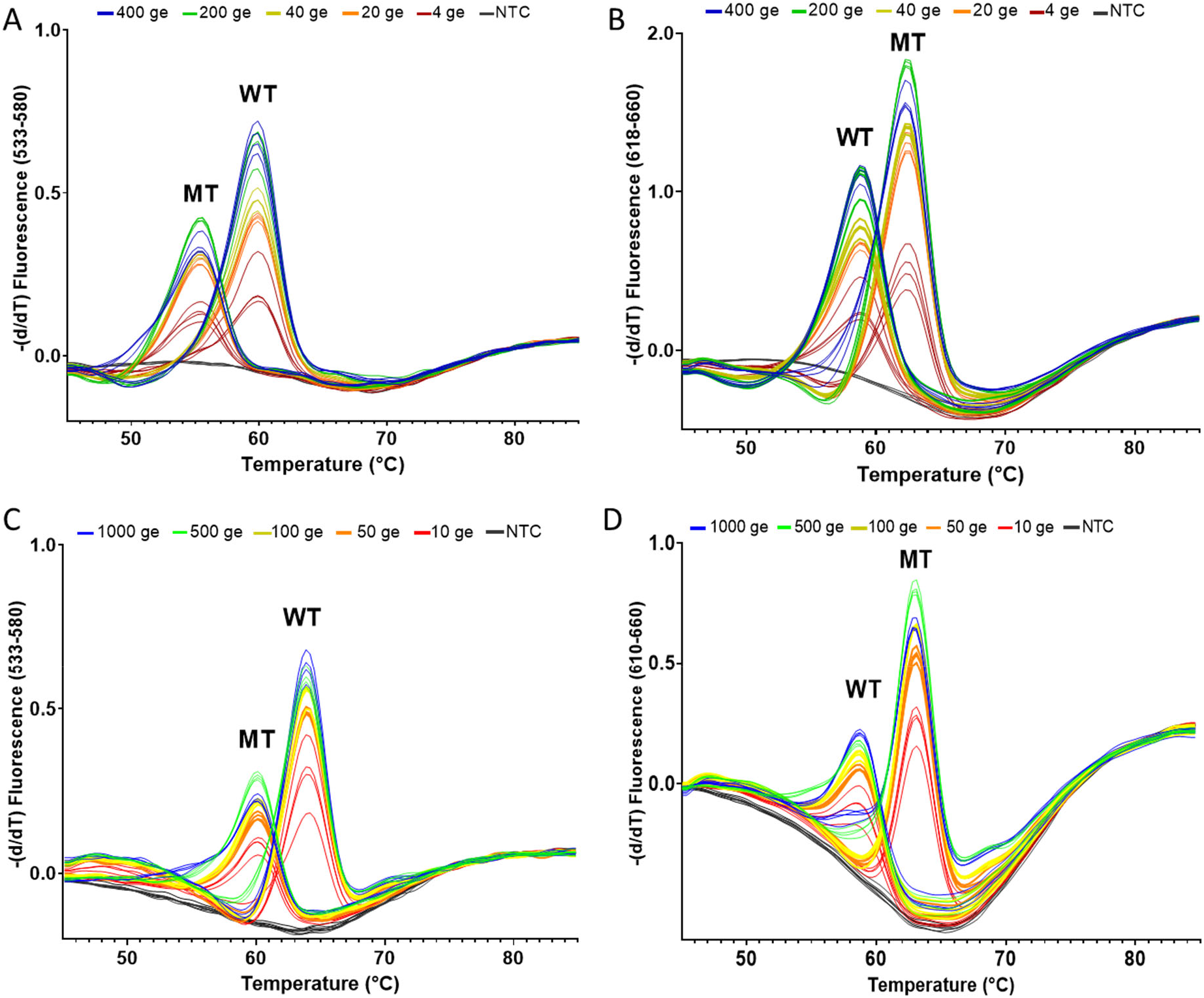
Analytical limit of detection and Tm values generated by the SMB-501 (A and B) and SMB-484 (C and D) assays tested against SARS-CoV-2 RNA. **A and C-** SARS-CoV-2 wildtype (WT) RNA; **B-** B.1.1.7 mutant (MT) RNA and **D**- B.351 mutant (MT) RNA at the indicated number of genomic equivalents (GEs).

### Tm code definition

The Tm values produced by both SMBs against the reference WT and the MT SARS-CoV-2 strains are listed in Table 2. The mean and standard deviations shown were derived from at least 4 replicates. When reference RNA was tested with the SMB 501-WT probe, WT-RNA produced a Tm of 59.8±0.4°C and MT N501Y RNA produced a Tm of 55.2±0.4°C. Similarly, when RNA was tested with the SMB 501-MT probe, WT-RNA produced a Tm of 58.2±1°C, and MT N501Y RNA produced a Tm of 62.25±0.6°C. Given that Tm values can vary slightly between clinical samples, we specified a two-temperature Tm code that identified either 501N or 501Y alleles within a Tm range of approximately 1.5-4 times the experimentally verified ±SD values for each probe Tm. Thus, the 501N (WT) Tm code was defined as a SMB-501-WT Tm of 59.8 ±1.5 °C and a SMB-501-MT Tm of 58.2 ±1.5°C; and a 501Y (MT) Tm code was defined as a SMB-501-WT Tm of 55.2 ±1.5 °C and a SMB-501-MT Tm of 62.25 ±1.5°C. Similarly, for SMB-484 assay, the reference Tm code for 484E (WT) was 64±0.1 (with SMB-484-WT probe) and 58.7±0.4 (with SMB-484-MT probe); and the reference Tm code for 484K (MT) was 59.7±0.5 (WT probe) and 63±0.6 (MT-probe). Any samples that failed to produce a Tm value for either of the SMBs or produced Tm values outside of the range defined for the Tm codes would have been defined as indeterminant and the assay repeated. Using this code definition, we retested our reference RNA samples 20 times, and the correct 501/484 allele was detected in each case, achieving an analytic sensitivity and specificity of 100%. These results clearly indicate that the combination of both SMB 501-WT and SMB 501-MT probes can specifically detect and differentiate the N501Y variants from the wild type strains with high confidence.

**Table 2.** Assay and Sanger sequencing results from COVID positive clinical samples.

| Samples | Date of 1st test<br>(dd/mm/yy) | PCR<br>Ct* | SMB-501 assay |  |  | SMB-484 assay |  |  | Confirmation by sequencing† |  |  |
| --- | --- | --- | --- | --- | --- | --- | --- | --- | --- | --- | --- |
|  |  |  | WT probe<br>(Cy3)<br>Tm (°C) | MT probe<br>(Cy5)<br>Tm (°C) | Id | WT probe<br>(Cy3)<br>Tm (°C) | MT probe<br>(Cy5)<br>Tm (°C) | Id | Wild<br>type | N501Y<br>mutant<br>(AAT-TAT) | E484K<br>mutant<br>(GAA-AAA) |
| WT-Reference |  |  | 59.8±0.4 | 58.2±1 | WT | 64±0.1 | 58.7±0.4 | WT | Yes | No | No |
| MT-Reference |  |  | 55.2±0.4 | 62.2±0.6 | MT | 59.7±0.5 | 63±0.6 | MT | No | Yes | No |
| SMBP-1 | Oct-Nov<br>2020 | 29.1 | 59.9 | 58.4 | WT | 63.9 | 58.2 | WT | Yes | No | No |
| SMBP-2 |  | 34.8 | NP | NP | Neg |  |  |  |  |  |  |
| SMBP-3 |  | 34.5 | NP | NP | Neg | 64.0 | 58.7 | WT |  |  |  |
| SMBP-4 |  | 35.4 | NP | NP | Neg |  |  |  |  |  |  |
| SMBP-5 |  | 39.2 | 59.5 | 58.2 | WT |  |  |  |  |  |  |
| SMBP-6 |  | 36.0 | 59.1 | 58.2 | WT |  |  |  | Yes | No | No |
| SMBP-7 |  | 33.7 | 58.9 | 58.1 | WT | 64.0 | NP | Ind |  |  |  |
| SMBP-8 |  | 36.8 | 59.5 | 58.2 | WT |  |  |  |  |  |  |
| SMBP-9 |  | 27.9 | 59.4 | 58.3 | WT |  |  |  |  |  |  |
| SMBP-10 | Jan 2021 | 29.4 | 59.3 | 58.3 | WT | 63.8 | 58.2 | WT | Yes | No | No |
| SMBP-11 |  | 39.1 | 59.9 | 58.6 | WT |  |  |  |  |  |  |
| SMBP-12 |  | 33.8 | 59.0 | 58.2 | WT | 64.3 | 58.8 | WT | Yes | No | No |
| SMBP-13 |  | 17.7 | 59.3 | 58.5 | WT | 64.2 | 58.9 | WT | Yes | No | No |
| SMBP-14 |  | 21.3 | 59.4 | 58.5 | WT | 64.2 | 58.8 | WT | Yes | No | No |
| SMBP-15 |  | 17.5 | 59.1 | 58.3 | WT | 64.0 | 58.7 | WT | Yes | No | No |
| SMBP-16 |  | 30.0 | 55.5 | 62.6 | MT | 64.2 | 58.5 | WT | No | Yes | No |
| SMBP-17 |  | 41.2 | NP | NP | Neg |  |  |  |  |  |  |
| SMBP-18 |  | 23.2 | 59.8 | 58.9 | WT | 64.0 | 58.7 | WT |  |  |  |
| SMBP-19 |  | 28.3 | 59.3 | 58.1 | WT | 63.8 | 58.2 | WT | Yes | No | No |
| SMBP-20 |  | 17.1 | 59.2 | 58.3 | WT | 64.0 | 58.8 | WT | Yes | No | No |
| SMBP-21 |  | 16.4 | 59.2 | 58.4 | WT | 64.2 | 58.9 | WT | Yes | No | No |
| SMBP-22 |  | 18.2 | 58.9 | 58.1 | WT |  |  |  | Yes | No | No |
| SMBP-23 |  | 21.1 | 58.9 | 58.1 | WT | 64.1 | 58.6 | WT | Yes | No | No |
| SMBP-24 |  | 39.0 | 59.5 | 58.2 | WT |  |  |  |  |  |  |
| SMBP-25 |  | 28.4 | 54.5 | 61.6 | MT | 64.1 | NP | Ind | No | Yes | No |
| SMBP-26 | Feb 2021 | 27.8 | 54.5 | 61.6 | MT | 63.9 | 58.3 | NP | No | Yes | No |
| SMBP-27 |  | 33.5 | 59.3 | 58.9 | WT | 60.1 | 63.0 | MT | Yes | No | Yes |
| SMBP-28 |  | 38.3 | 59.7 | 58.5 | WT | 64.1 | NP | Ind | ND | ND | ND |
| SMBP-29 |  | 18.1 | 58.9 | 58.1 | WT | 64.1 | 58.8 | WT | Yes | No | No |
| SMBP-30 |  | 27.8 | 59.2 | 58.2 | WT |  |  |  |  |  |  |
| SMBP-31<sup>a</sup> |  | 17.6 | 55.1 | 61.9 | MT | 60.4 | 63.3 | MT | No | Yes | Yes |
| SMBP-32<sup>a</sup> |  | 17.4 | 55.1 | 61.9 | MT | 64.0 | 58.7 | WT | No | Yes | No |
| SMBP-33 |  | 32.8 | 55.0 | 62.1 | MT |  |  |  |  |  |  |
| SMBP-34 |  | 25.2 | 55.5 | 62.7 | MT | 60.2 | 63.3 | MT | No | Yes | Yes |
| SMBP-35 |  | 18.4 | 55.6 | 62.5 | MT | 60.5 | 63.4 | MT | No | Yes | Yes |
| SMBP-36 |  | 18.8 | 54.9 | 61.9 | MT |  |  |  | No | Yes | No |
| SMBP-37 |  | 28.4 | 54.9 | 62.2 | MT |  |  |  |  |  |  |
| SMBP-38 |  | 16.4 | 59.8 | 58.5 | WT | 64.1 | 58.6 | WT |  |  |  |
| SMBP-39 |  | 32.2 | NP | NP | Neg |  |  |  |  |  |  |
| SMBP-40 |  | 32.3 | 58.9 | 57.8 | WT |  |  |  | Yes | No | No |
| SMBP-41 | March 2021 | 32.6 | 55.4 | 62.6 | MT | 63.9 | 58.3 | WT | No | Yes | No |
| SMBP-42 |  | 36.8 | 59.7 | 58.4 | WT |  |  |  |  |  |  |
| SMBP-43 |  | 30.1 | 59.8 | 58.9 | WT |  |  |  | Yes | No | No |
| SMBP-44 |  | 33.3 | NP | NP | Inv |  |  |  |  |  |  |
| SMBP-45 |  | 34.6 | NP | 61.3 | Ind |  |  |  |  |  |  |
| SMBP-46 |  | 23.3 | 59.2 | 58.285 | WT | 64.0 | 58.6 | WT | Yes | No | No |

IC-internal control; WT-wildtype; MT-Mutant; NP- No Tm peak; Neg-Negative; ND- not Inv-Invalid; Ind-Indeterminate.
\*Xpert Xpress SARS-CoV-2 or Xpert Xpress SARS-CoV-2/Flu/RSV tests
<sup>†</sup>Representative strains were sequenced. <sup>a</sup>sequencing was repeated twice to confirm the identified mutations.
Blocked grey cells- Corresponding samples that are not tested by the SMB-484 assay or sequencing.

### Validation with patient samples

A total of 76 patient samples, 46 confirmed COVID-19 positive and 30 confirmed COVID-19 negative, were tested using the SMB-501 assay. A subset of 45 of these same samples (26 COVID-19 positive and 19 COVID negative) were additionally validated with the SMB-484 assay. None of the COVID-19 negative samples produced any measurable Tm values from either of the assays, yielding a specificity of 100%. Positive samples were selected based on a wide range of N2-Ct values between 16 through 41 based on the Xpert Xpress SARS-CoV-2 assay. As shown in Table 2, 40/45 (89%) of the COVID-19 positive samples produced measurable Tm values for both SMB probes in the SMB-501 assay, in at least one of the two replicates. Two samples were identified as indeterminate based on the inability of either the WT or MT probe to generate a measurable Tm and/or due to the presence of a failed internal control. There were also 5 assays from previously positive clinical samples, which generated negative results in that neither probe produced a measurable Tm. All the negative assays had shown an initial Xpert Ct values ≥35. Although, many of the samples with Xpert Ct values as late as 39 could still be detected by our assay, we found that the samples with a Ct ≥35 using the Xpert assay, yielded relatively stunted melt peak heights of ≤0.3 with the WT-SMB and ≤0.1 with MT-SMB similar to that observed when testing 4 GE in our limit of detection studies, which is indicative of very low viral loads in these samples. However, the Tm could still be identified for at least one of the 501 SMB probes in all samples, except the negatives. Overall, 27/45 (60%) of the COVID-19 positive samples had SMB 501-WT and SMB 501-MT Tm values consistent with the WT N501 allele and 11/45 (24.4%) of the samples had Tm values consistent with mutant Y501 allele within ± 1-standard deviation of our reference standards (Table 1, Fig. 2). Twelve representative WT and mutant samples as predicted by the SMB assay, underwent Sanger sequencing of the PCR products for confirmation of the PCR results. In all cases, the WT or mutant sequences identified by the SMB N501Y assay were confirmed by the sequencing result. Using the confirmed sequencing results as a gold standard, 4/4 of the wild type clinical samples were detected as WT by the SMB N501Y assay, and 8/8 N501Y mutant results were detected as mutant by the assay, demonstrating a clinical sensitivity and specificity of 100%. Similarly, with the SMB-484 assay, 19/26 (73%) of the samples tested were wild type and 4/26 (15.3%) were variants. Out of the 4 variants, 4/4 were sequenced and confirmed to harbor the G>A mutation in the 484^th^ codon, yielding an assay specificity of 100% compared to sequencing. It should be noted that one of the specimens (SMBP-27) contained a E484K mutation but was found to have a WT sequence at codon 501. Considering that the major three VOCs contain either N501Y alone (B.1.1.7) or both N501Y and E484K mutations (B.1.351 and P.1), the presence of this single 484K mutant might indicate that this sample contains the variant of interest B.1.525/B.1.526, which originated in New York City. Thus, the combination of our SMB 501 and SMB-484 assays should help capture most of the variants in circulation. We also observed that the frequency of N501Y and E484K variants increased substantially between the clinical samples obtained in October 2020 and the samples obtained in February and March 2021 (Fig. 3).

**Fig. 2.**
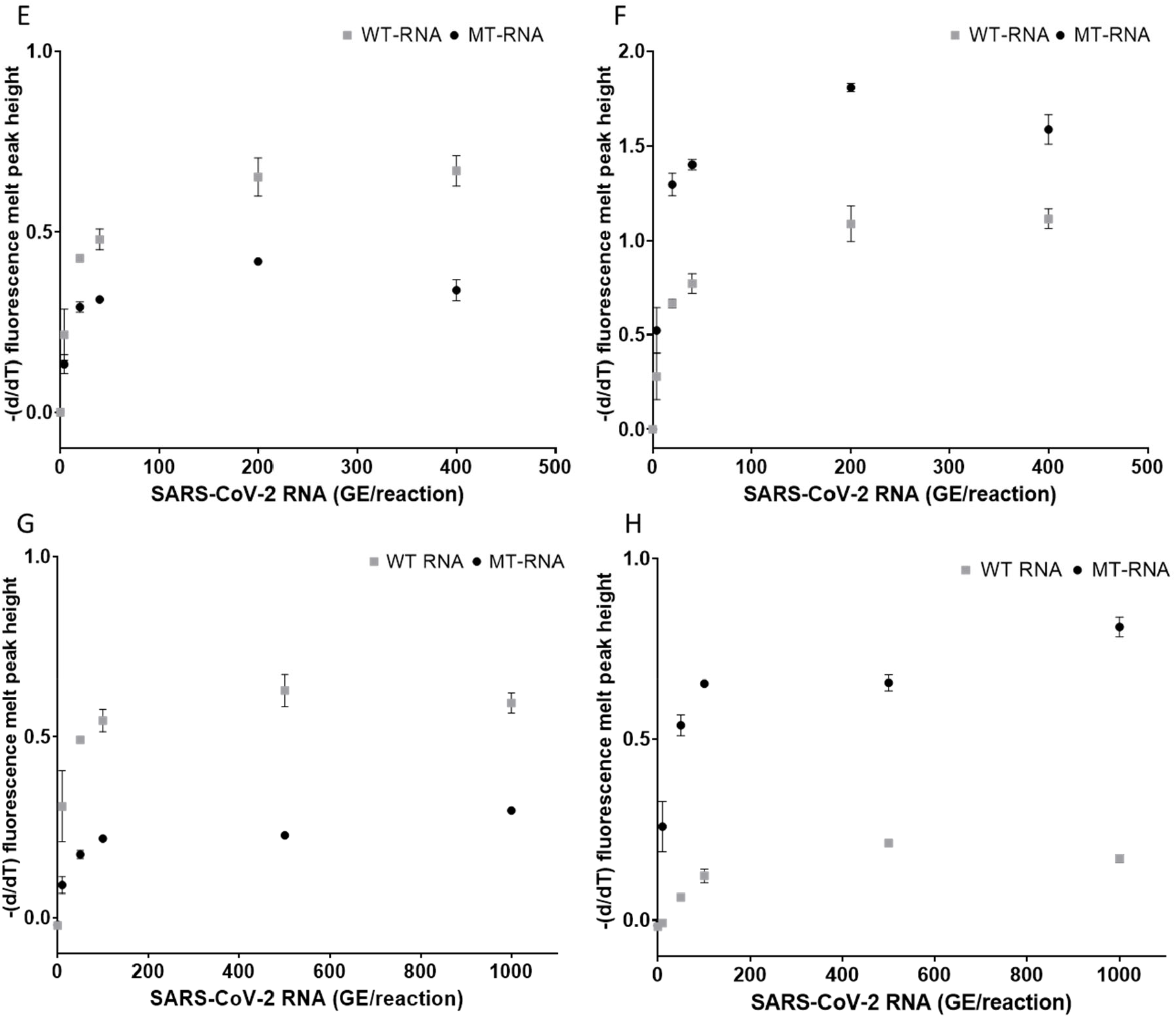
The effect of target concentration on the melt peak height generated by the SMB-501 (A and B) and SMB-484 (C and D) assays tested against SARS-CoV-2 RNA. **A and C-** SARS-CoV-2 wildtype (WT) RNA; **B-** B.1.1.7 mutant (MT) RNA and **D**- B.351 mutant (MT) RNA at the indicated number of genomic equivalents (GEs).

**Fig. 3.**
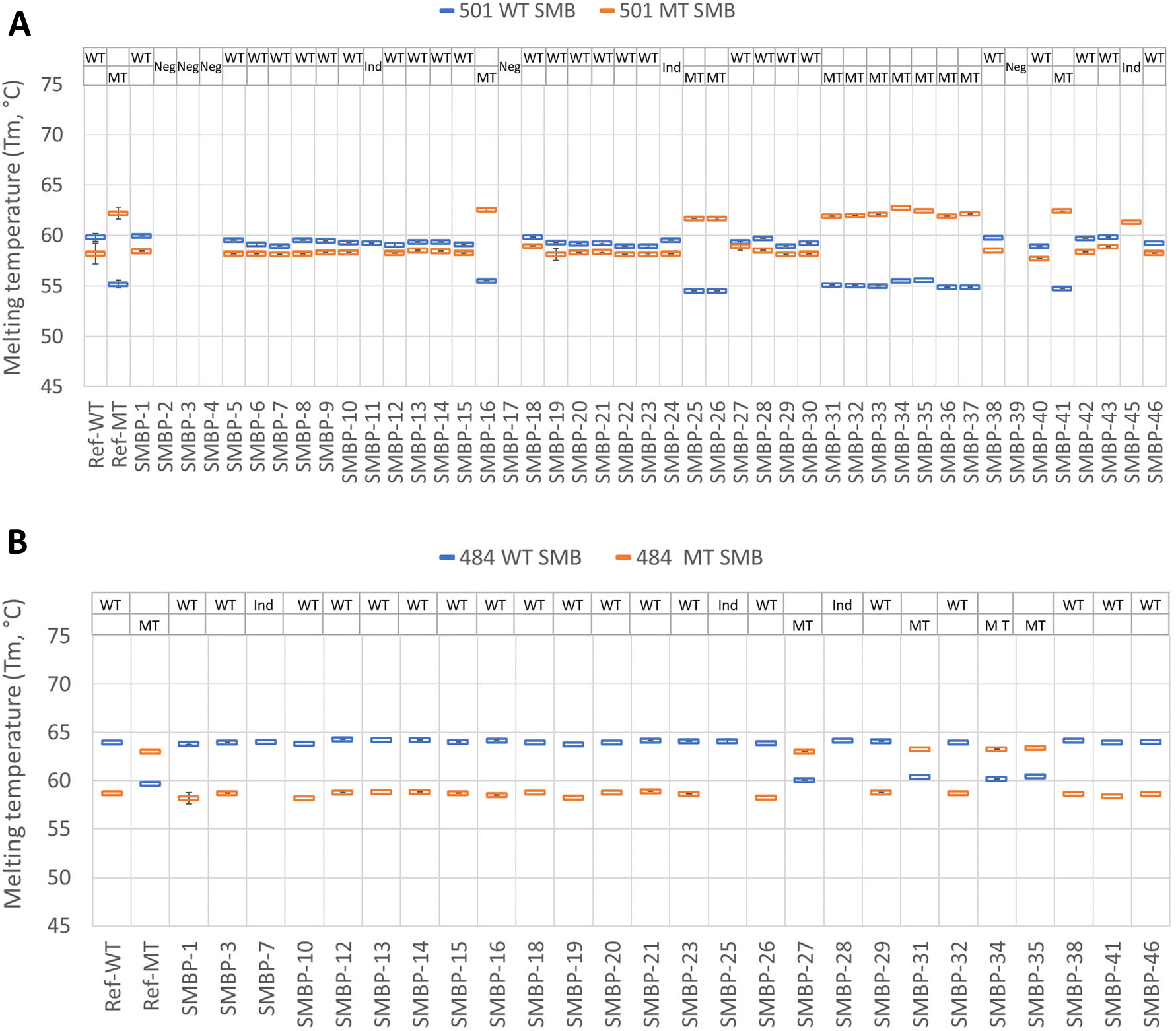
Sloppy molecular beacon (SMB) Tm profile of positive clinical nasopharyngeal (NP) samples tested using the SMB-501 and SMB-484 Tm assay. Tm signatures consisting of the Tm values for both the WT probe (blue) and MT probe (orange) are shown for the SMB-501 assay **(A)** and the SMB-484 assay **(B)**. Ref WT indicates the Tm profile of the reference, WT SARS-CoV-2 strain and Ref MT indicates the TM profile of the reference MT SARS-CoV-2 B.1.1.7 strain (A) and B.351 (B). SMBP1 – SMBP46 indicate that Tm profiles of the 46 COVID positive clinical samples tested in this study with SMB-501 assay and the 26 COVID positive samples tested by SMB-484 assay. Error bars show +/-one standard deviation.

**Fig.4.**
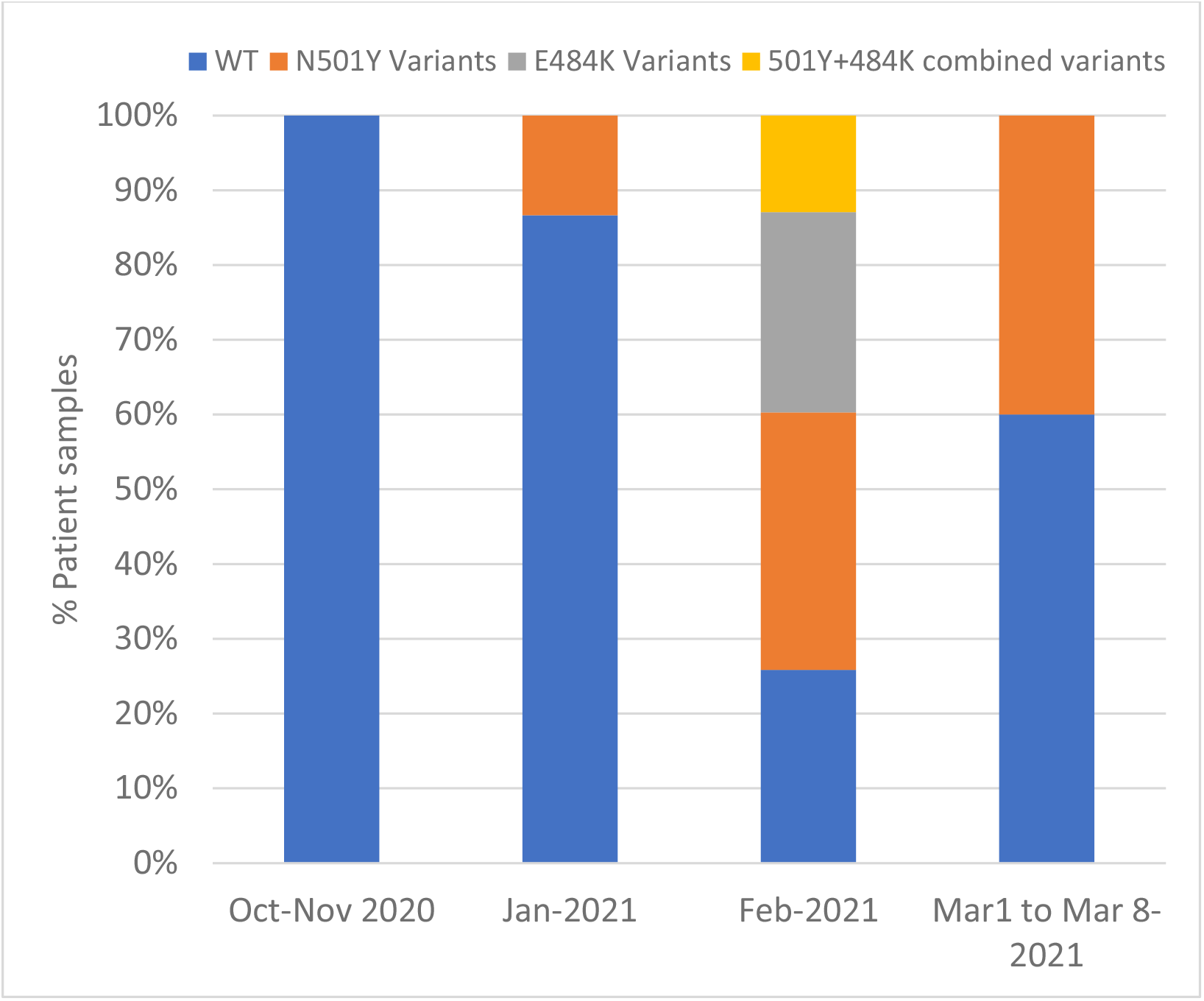
Prevalence of N501Y variant strains among the tested sample set over time. The proportion of N501Y and E484K variants is shown for samples obtained during the periods of October-November (n=9 tested with SMB-501/n=3 tested with SMB-484), January (n=16/n=12), February (n=15/n=9) and the first week of March (n=6/n=2). No samples from December were tested in our study.

## Discussion

The emergence of SARS-CoV-2 VOC with the potential for increased transmission, disease severity, and resistance to vaccine induced immunity is of grave concern (14, 28). A simple screening assay to monitor the emergence and spread of these strains may be helpful for implementing public health strategies to counter these and future strains. Our study demonstrates that our assay is simple, rapid, and sensitive and specific for detecting key variant-identifying mutations using a high-throughput PCR assay platform. Thus, this assay has the potential for relatively inexpensive high throughput testing for rapid identification of N501Y, E484K variants. In designing this assay, we took advantage of the fact that the N501Y and E484K mutations are common in the major SARS-CoV-2 variants. Since this mutation also appears to be responsible for the increased infectivity and possibly the other adverse manifestations of these strains (4), assays which detect this mutation may also prove useful to detect any future strain that evolves to have increased transmission potential.

Our assay is the first to our knowledge that uses post PCR Tm based analysis to detect and differentiate SARS-CoV-2 variants using SMB probes demonstrated in clinical samples. This assay format has the benefit of producing a measurable Tm result irrespective of whether the SMB probe is fully complementary to its target nucleic acid sequence. Instead of detecting a mutation by either producing or not producing a signal, SMBs detect mutations by producing a Tm shift. Failure to produce a Tm signal indicates an invalid assay rather than the absence of a mutation. The robustness of our assay is further increased by our use of two different SMBs, one complementary to the WT sequence and one complementary to the mutant sequence. The pattern of Tm values or “Tm signature” produced by the combined Tm values of each SMB probe provides an unequivocal identification of a WT or MT sequence. We have also shown that Tm signatures can be used to detect mixtures of mutant and WT sequences and to identify numerous mutations present in an assay’s target region (29). Thus, we expect that our assay should continue to be able to identify N501Y and E484K variants even if additional mutations develop near this primary mutation within the probe-footprint region once the specific Tm signatures of each new genotype are characterized. Our assay is meant to be a screening assay which will identify samples likely to contain SARS-CoV-2 variants of concern. Although in this study, we have proposed to use the combination of two probes per assay for increased accuracy, we expect that most SARS CoV-2 variants can be identified using only the MT-probe alone, and future versions of our assay are likely to use a single SMB to identify mutations that are less critical than N501Y and E484K. We suggest that a useful public health strategy would be to screen COVID-19 positive samples in near real time with our assay, and then to perform genomic sequencing on a subset of screen-positive samples. This sequencing would confirm the presence of the expected variants and in some cases lead to the discovery of new variants. Ongoing sequencing of a subset of all SARS-CoV-2 samples will also be required to identify completely novel variants or to investigate the epidemiology and clinical characteristics of variants such as those recently reported in the United States. Fortunately, our assay is easily extensible and additional tests for new key mutations can be added in a modular format to a screening panel when new mutations associated with critical new variants are discovered. In fact, it is our intention to continuously update this assay until our work is superseded by a better approach or much more widespread genome sequencing becomes commonplace. Key updates will also be posted on a preprint journal. In the meantime, we hope that our current test will help increase surveillance and potentially help control the spread of the new emerging variants of concern.

## Data Availability

The assay published here along with the primers and probes are made available freely for the readers

## Acknowledgements

This study was funded by the National Institute of Allergy and Infectious Diseases of the National Institutes of Health under award number R01 AI131617. The following reagent was deposited by the Centers for Disease Control and Prevention and obtained through BEI Resources, NIAID, NIH: Genomic RNA from SARS-Related Coronavirus 2, Isolate USA-WA1/2020, NR-52285.The following reagent was obtained through BEI Resources, NIAID, NIH: Severe Acute Respiratory Syndrome-Related Coronavirus 2, Isolate hCoV-19/England/204820464/20200, NR-54000, contributed by Bassam Hallis. The authors wish to thank all the laboratories that contribute sequence data to the GISAID EpiCoV database. A GISAID acknowledgment table reporting the geographic origin and contributions of genomes analyzed in this study is attached as a supplementary Table 1S.

D.A. receives research support and royalty payments from Cepheid, which sells the Xpert Xpress SARS-CoV-2 and Xpert Xpress SARS-CoV-2/Flu/RSV tests., S.C, is an employee of Cepheid and R.J. is a paid consultant for Cepheid.

